# Machine Learning with Objective Serum Markers and Algorithmic Deep Learning CT Scan Analysis Predicts Brain Death Due to Trauma and Hypoxia

**DOI:** 10.1101/2021.02.13.21251369

**Authors:** Daniel Rafter, Brett Sterk, Zhuliu Li, Tory Schaaf, Rui Kuang, Michael Koller, Uzma Samadani

## Abstract

**Background:** Prediction of which traumatic and hypoxic brain injuries will progress to brain death is currently based predominantly on history, physical examination and radiographic findings. We investigated how accurately purely objective measures including three neurologic serum markers and algorithmic CT (computed tomograph) scan analysis can predict brain death and differentiate its etiology.

**Methods:** This prospective observational study enrolled 51 isolated trauma subjects and 19 clinically brain dead subjects that were further divided by mechanism of injury into high-velocity trauma with presumed diffuse axonal injury, cardiopulmonary/respiratory arrest, and found down groups. Levels of glial fibrillary acidic protein (GFAP), ubiquitin carboxy-terminal hydrolase L1 (UCH-L1), and S100 Calcium-Binding Protein B (S100B) were compared, and algorithmic analysis of CT scan imaging was performed using an open-source deep learning software (BLAST-CT).

**Findings:** The prognostic value of various biomarker combinations in identifying subjects progressing to brain death was assessed using machine learning. Prediction accuracy for GFAP, UCH-L1, and S100B and algorithmic CT analysis in combination to predict brain death from among all other cohorts yielded an area under the receiver operator curve of 0.98 versus all other brain injury subjects. This model was also able to distinguish brain death attributed to cardiopulmonary/respiratory arrest from combined non-trauma controls and diffuse axonal injury with an area under the curve of 0.99.

**Interpretation:** Serum concentrations of GFAP, UCH-L1, and S100B measured within 32 hours of traumatic and hypoxic brain injury (cardiopulmonary/respiratory arrest) and algorithmic CT analysis have utility in prognosticating brain death and predicting its mechanism of injury as either hypoxic or traumatic/unknown (diffuse axonal injury/found down).

**Funding:** Abbott Labs and the Minnesota State Office of Higher Education.

## INTRODUCTION

As many as half of all trauma patients present with some level of intoxication, and a high percentage have comorbidities which can potentially obfuscate early detection of severe acute injuries. Obtaining a patient’s history, physical examination and computed tomograph (CT) are the current gold standard for assessment in the acute phase of brain injury (Greer et al., 2020). The history however, is sometimes unavailable due to patient factors or unreliable due to clinician bias. The physical examination can be confounded by multiple factors, and the radiologist interpretation of CT scan is not quantifiable or standardized. Thus there may be variability in patient outcomes as predicted by these subjective measures (Lingsma et al., 2010).

Severe brain injury is a leading cause of disability worldwide (Faul and Coronado, 2015) and its mechanistic heterogeneity has proven to be a barrier for advancing both diagnostic and prognostic technologies. It is estimated that 39% of patients with a severe traumatic or atraumatic brain injury (Rosenfeld et al., 2012) progress to brain death within their initial admission. The mechanistic heterogeneity of severe brain injury implies that some injuries may potentially be more reversible than others, but the point at which injury becomes irreversible, and differentiating among the pathologies to identify any reversible processes is currently challenging. Two mechanistically dichotomous pathologies that may potentially advance to brain death are traumatic diffuse axonal injury and atraumatic anoxic or hypoxic brain injury. Severe edema, hypoxia, or infarction are associated with these pathologies and contribute to brain death, resultant edema, loss of blod flow and neuronal death. A diffuse axonal injury results from high velocity shearing forces at the gray-white matter interface, particularly in the corpus callosum and brainstem (Mesfin et al., 2020). Anoxia results after as little as 20 seconds of circulatory arrest depletes neuronal oxygen stores resulting in loss of consciousness. Continued insufficient circulatory support for 10 minutes is generally lethal. Radiographic signs of hypoxic injury are frequently undetectable in the acute setting, and sequelae of structural disruption including global edema and diffusion weighted changes may not be evident until days after the original insult making accurate prognostication difficult (Nguyen et al., 2018). As such, there is a strong need for improved prognostic measures of brain death in diffuse axonal injuries, anoxic brain injury due to cardiac or respiratory arrest, and brain injury of unknown origin in order to make informed clinical decisions about withdrawing life support, avoiding futile interventions, allocating resources and monitoring progression and therapeutic responses (Wang et al., 2018).

Serum biomarkers offer potential to aid in objective measurement and prognostication of brain injury (Wang et al., 2018), and have been examined in various brain injury cohorts including brain death (Mahan et al., 2019). In particular, glial fibrillary acidic protein (GFAP), the predominant filament protein in the astrocytic skeleton has demonstrated potential as a marker of astrocytic injury (Mahan et al., 2019), and is markedly upregulated during astrogliosis, a process in which astrocytes increase in size and number in response to traumatic and anoxic injury (Egea-Guerrero et al., 2013). Ubiquitin carboxy-terminal hydrolase L1 (UCH-L1) is a neuron-specific protein, known for its role in the addition and removal of ubiquitin from proteins (Papa et al., 2016). Much like GFAP, UCH-L1 is elevated in the serum following a traumatic brain injury and is predictive of injury severity and presence of intracranial lesions on head CT (Diaz-Arrastia et al., 2014). S100B is a well-studied biomarker that functions in astrocytes as a major homeostatic calcium-binding protein and is released into the serum after brain injury (Rodriguez-Rodriguez et al., 2016). One study investigated S100B in both severe traumatic brain injured and cardiac arrest patients, finding that concentrations measured at 12 hours post-admission were comparable between traumatic brain injured and cardiac arrest groups, as well as predictive of unfavorable neurologic outcome (Mussack et al., 2002).

The aim of this study is to explore the potential role of serum biomarkers and algorithmic open-source deep-learning CT scan analysis (Mahan et al., 2019) in predicting brain death in both non-traumatic and traumatic brain injuries. Our goal is to use machine learning to develop a brain injury stratification model based on algorithmic CT scan analysis and concentrations of three serum markers GFAP, UCH-L1, and S100B taken within 32 hours of hospital admission. The 32 hour time point was chosen based on previous work demonstrating that serum biomarker concentrations during this time frame can be used to accurately predict CT positive and CT negative scans from patients with traumatic brain injuries (Mahan et al., 2019). This model will predict those at risk for brain death from among patients who suffered a traumatic or atraumatic brain injury. Evaluation of these objective measures may be useful in classifying the pathophysiology of injury as due to cardiac/respiratory arrest (CA/RA), unknown etiology/found down (FD), or diffuse axonal injury (DAI).

## MATERIALS &METHODS

### Study Design

Institutional Review Board approval (HSR #15-4079) was obtained for prospective enrollment of patients who presented to the emergency department of a level-1 trauma center between May 2016 and April 2017. Subjects were consented if they were able to pass the Galveston Orientation and Amnesia Test (GOAT) and met criteria for providing informed consent. If they were not able to provide informed consent initially, a legally authorized representative was consented and the subject was reconsented once able. Only in situations where the subject expired in the hospital and family or a legally authorized representative could not be contacted was informed consent waived. Informed consent or waiver of consent was obtained prior to data inclusion. Specific information regarding study setting, population, determination of key outcome measures, neuroimaging techniques, classification, blood collection and processing, and machine learning are included in the supplemental materials.

### Machine Learning

Support Vector Machine (SVM) (Corinna Cortes, 1995) was utilized for the classification of the patient samples in our prediction tasks. Among the supervised learning methods, SVM generalizes well on new test data, and the decision function of SVM is simply defined by a subset of training samples without requiring training with big data (Supplemental Methods).

### Leave-one-out cross-validation and parameter tuning

We perform leave-one-out cross-validation to evaluate the prediction performance of SVM. In the evaluation, we hold out one sample at a time as the test data and treat the rest samples as training data to obtain the SVM model for classifying the held-out sample in the test data. We repeat the procedure on every sample and report the overall test accuracy in Table 3 and Table 4.

**Table 1.** Study demographics cohorts.

| Data Point |  | Brain Death | CT (+)<br>TBI | CT (-)<br>TBI | Non-Trauma | Controls |
| --- | --- | --- | --- | --- | --- | --- |
| N |  | 19 | 21 | 30 | 15 | 41 |
| Male \ Female |  | 13 : 6 | 15 : 6 | 21 : 9 | 8 : 7 | 11 : 30 |
|  |  | P-value (chi squared) | 0.84 | 0.004 | 0.029 | 0.0000000001 |
| Age (mean, range) |  | 58.6, 24 - 87 | 56.2, 31 - 85 | 50.4, 4 - 95 | 53.8, 18 - 86 | 37.1, 16 - 65 |
|  |  | P-value | 0.69 | 0.23 | 0.42 | 0.000004 |
| LOC | None | 1 (5%) | 3 (14%) | 15 (50%) | 5 (33%) | 41 |
|  | 0 - 30 mins | 2 (11%) | 6 (29%) | 8 (27%) | 1 (7%) | - |
|  | 30 mins - 24 hrs | 0 (0%) | 2 (10%) | 0 (0%) | 1 (7%) | - |
|  | > 24 hrs | 14 (74%) | 0 (0%) | 0 (0%) | 0 (0%) | - |
|  | Unknown | 2 (11%) | 10 (48%) | 7 (23%) | 8 (53%) | - |
| GCS | 13 - 15 | 1 (5%) | 13 (62%) | 28 (93%) | 6 (40%) | 41 |
|  | 9 -12 | 0 (0%) | 3 (14%) | 1 (3%) | 1 (7%) | - |
|  | 8 or less | 18 (95%) | 5 (24%) | 1 (3%) | 8 (53%) | - |
| Percent with clinical CT |  | 95% | 100% | 100% | 100% | 0 % |
| Marshall Score (mean, range) |  | 3.1, 1 - 6 | 2.14, 2 - 4 | 1 | 2.47, 1 - 6 | - |
| Length of Stay | 0 - 48 hrs | 5 | 7 | 27 | 0 | - |
|  | 49 - 96 hrs | 4 | 5 | 2 | 3 | - |
|  | > 96 hrs | 10 | 9 | 1 | 12 | - |
| Discharge Location | Home | 0 | 13 | 29 | 4 | - |
|  | Rehab | 0 | 8 | 1 | 11 | - |
|  | Morgue | 19 | 0 | 0 | 0 | - |

**Table 2.**
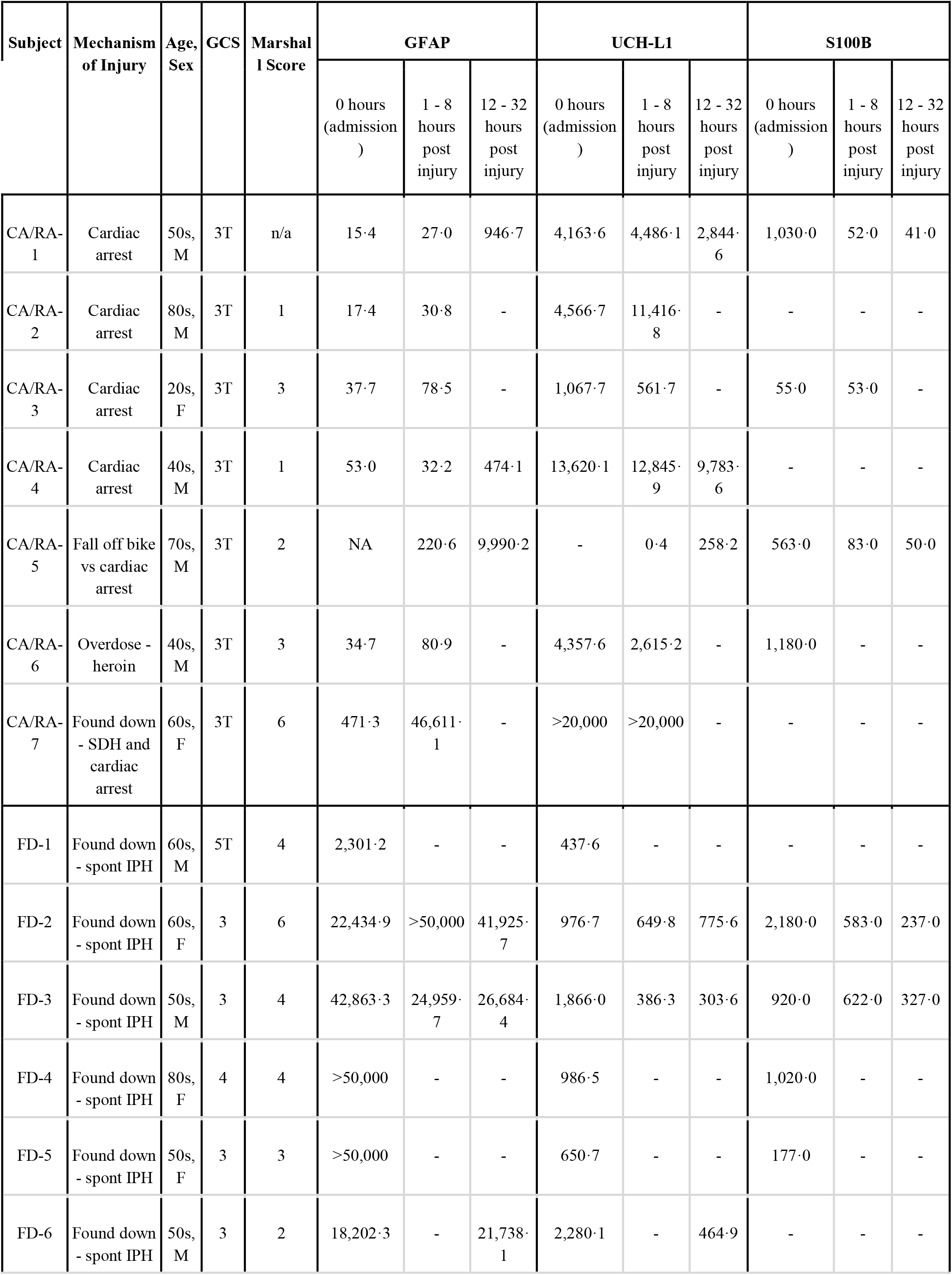
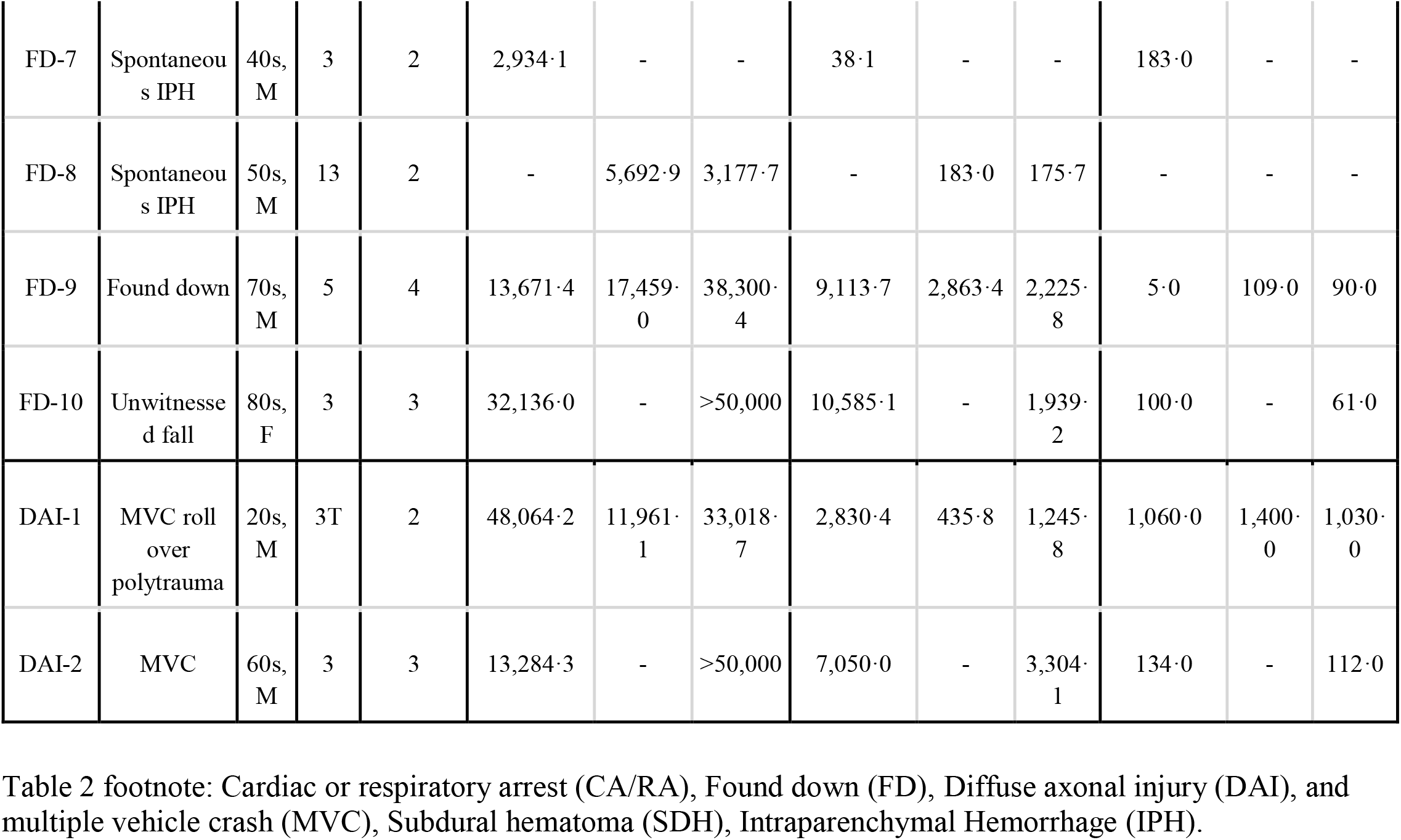
Concentrations (pg/mL) of GFAP, UCH-L1, and S100B from brain death cohorts shown at each blood draw time point.

| Subject | Mechanism of Injury | Age, Sex | GCS | Marshall Score | GFAP |  |  | UCH-L1 |  |  | S100B |  |  |
| --- | --- | --- | --- | --- | --- | --- | --- | --- | --- | --- | --- | --- | --- |
|  |  |  |  |  | 0 hours (admission ) | 1 - 8 hours post injury | 12 - 32 hours post injury | 0 hours (admission ) | 1 - 8 hours post injury | 12 - 32 hours post injury | 0 hours (admission ) | 1 - 8 hours post injury | 12 - 32 hours post injury |
| CA/RA-1 | Cardiac arrest | 50s, M | 3T | n/a | 15.4 | 27.0 | 946.7 | 4,163.6 | 4,486.1 | 2,844.6 | 1,030.0 | 52.0 | 41.0 |
| CA/RA-2 | Cardiac arrest | 80s, M | 3T | 1 | 17.4 | 30.8 | - | 4,566.7 | 11,416.8 | - | - | - | - |
| CA/RA-3 | Cardiac arrest | 20s, F | 3T | 3 | 37.7 | 78.5 | - | 1,067.7 | 561.7 | - | 55.0 | 53.0 | - |
| CA/RA-4 | Cardiac arrest | 40s, M | 3T | 1 | 53.0 | 32.2 | 474.1 | 13,620.1 | 12,845.9 | 9,783.6 | - | - | - |
| CA/RA-5 | Fall off bike vs cardiac arrest | 70s, M | 3T | 2 | NA | 220.6 | 9,990.2 | - | 0.4 | 258.2 | 563.0 | 83.0 | 50.0 |
| CA/RA-6 | Overdose - heroin | 40s, M | 3T | 3 | 34.7 | 80.9 | - | 4,357.6 | 2,615.2 | - | 1,180.0 | - | - |
| CA/RA-7 | Found down - SDH and cardiac arrest | 60s, F | 3T | 6 | 471.3 | 46,611.1 | - | >20,000 | >20,000 | - | - | - | - |
| FD-1 | Found down - spont IPH | 60s, M | 5T | 4 | 2,301.2 | - | - | 437.6 | - | - | - | - | - |
| FD-2 | Found down - spont IPH | 60s, F | 3 | 6 | 22,434.9 | >50,000 | 41,925.7 | 976.7 | 649.8 | 775.6 | 2,180.0 | 583.0 | 237.0 |
| FD-3 | Found down - spont IPH | 50s, M | 3 | 4 | 42,863.3 | 24,959.7 | 26,684.4 | 1,866.0 | 386.3 | 303.6 | 920.0 | 622.0 | 327.0 |
| FD-4 | Found down - spont IPH | 80s, F | 4 | 4 | >50,000 | - | - | 986.5 | - | - | 1,020.0 | - | - |
| FD-5 | Found down - spont IPH | 50s, F | 3 | 3 | >50,000 | - | - | 650.7 | - | - | 177.0 | - | - |
| FD-6 | Found down - spont IPH | 50s, M | 3 | 2 | 18,202.3 | - | 21,738.1 | 2,280.1 | - | 464.9 | - | - | - |
| FD-7 | Spontaneous IPH | 40s, M | 3 | 2 | 2,934·1 | - | - | 38·1 | - | - | 183·0 | - | - |
| FD-8 | Spontaneous IPH | 50s, M | 13 | 2 | - | 5,692·9 | 3,177·7 | - | 183·0 | 175·7 | - | - | - |
| FD-9 | Found down | 70s, M | 5 | 4 | 13,671·4 | 17,459·0 | 38,300·4 | 9,113·7 | 2,863·4 | 2,225·8 | 5·0 | 109·0 | 90·0 |
| FD-10 | Unwitnessed fall | 80s, F | 3 | 3 | 32,136·0 | - | >50,000 | 10,585·1 | - | 1,939·2 | 100·0 | - | 61·0 |
| DAI-1 | MVC roll over polytrauma | 20s, M | 3T | 2 | 48,064·2 | 11,961·1 | 33,018·7 | 2,830·4 | 435·8 | 1,245·8 | 1,060·0 | 1,400·0 | 1,030·0 |
| DAI-2 | MVC | 60s, M | 3 | 3 | 13,284·3 | - | >50,000 | 7,050·0 | - | 3,304·1 | 134·0 | - | 112·0 |

**Table 3:** Characterization of brain death biomarkers.

| Biomarker |  | Brain Death | CT (+) TBI | CT (-) TBI | Non-Trauma | Controls |
| --- | --- | --- | --- | --- | --- | --- |
| GFAP | samples (n) | 40 | 46 | 49 | 27 | 41 |
|  | mean, stdev | 18,248.3, 19,254.6 | 2,871.7, - 4,992.5 | 134.8, 263.7 | 2,489.5, 6,104.3 | 13.5, 14.4 |
|  | range | 15.4 - 50,000 | 33.2 - 22,178.8 | 4.7, 1,426.1 | 3.3 - 27,985 | 0.6 - 84.5 |
|  | t test p value vs brain death | - | 0.0000133 | 0.0000006 | 0.0000135 | 0.0000005 |
| UHC-L1 | samples (n) | 40 | 46 | 49 | 27 | 41 |
|  | mean, stdev | 4,108.9, 5,287.1 | 637.9, 685.8 | 381.2, 639.6 | 433.0, 718.2 | 90.6, 70.9 |
|  | range | 0.4 - 20,000 | 4.2 - 2918.5 | 56 - 4,347.2 | 45.4 - 3,125.3 | 0 - 370.5 |
|  | t test p value vs brain death | - | 0.0001830 | 0.0000709 | 0.0000910 | 0.0000230 |
| S100B | samples (n) | 28 | 30 | 24 | 17 | 38 |
|  | mean, stdev | 480.6 - 548.6 | 403.6, 883.0 | 551.5, 1,316.6 | 275.2, 468.4 | 74.3, 42.1 |
|  | range | 5 - 2,180 | 16 - 4,280 | 25 - 6450 | 23 - 1,840 | 17 -207 |
|  | t test p value vs brain death | - | 0.6940469 | 0.8071675 | 0.2060105 | 0.0005538 |

**Table 4.** SVM prediction accuracy from two serum biomarkers (GFAP and UCH-L1) and inclusion of multimodal prediction with Blast-CT output.

| Comparison Groups<br>n = 203 samples | GFAP, UCH-L1 |  | GFAP, UCH-L1, Blast-CT |  |
| --- | --- | --- | --- | --- |
|  | AUC | AP | AUC | AP |
| **BD vs. CTL, NT, CTn, CTp** | 0.95 | 0.85 | 0.95 | 0.86 |
| **BD vs. NT, CTn, CTp** | 0.94 | 0.85 | 0.95 | 0.88 |
| **BD vs. NT, CTp** | 0.91 | 0.88 | 0.93 | 0.90 |
| **CA vs. FD, DAI** | 0.96 | 0.96 | 0.99 | 0.98 |

The parameters and 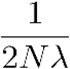 are chosen from (0.1, 0.5, 1, 5, 10) by grid searching.

### Measurements of the Prediction Accuracy

We comprehensively measured the classification accuracy using the Area Under the receiver operator Curve (AUC), and Average Precision (AP) (Fawcett, 2006; Zhang E., 2009). Denoting the number of true positives, true negatives, false positives and false negatives as TP, TN, FP, and FN respectively, the true positive rate (TPR), false positive rate (FPR) and precision (PR) are defined as:

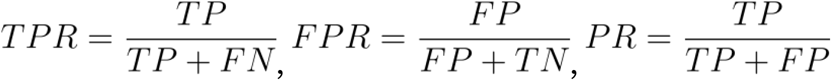

- Area Under the receiver operator Curve (AUC) measures the area under the receiver operating characteristic curve which plots the true positive rate (TPR) and false positive rate (FPR) at different classification thresholds. Thus, the AUC represents the probability that a randomly chosen positive sample receives a higher score than a randomly chosen negative sample.
- Precision@K measures the fraction of detected positive samples among the top-K predictions of the machine learning model, i.e., if there are N true positives among the top-K predictions then Precision@K is defined as

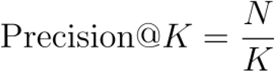
- Average Precision (AP) measures the mean of the precision values after each positive sample is detected, which is defined as

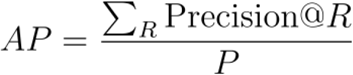

where *R* is the rank of each positive sample, and *P* is the total number of positive samples in the data. A high average precision score indicates that the SVM classifier can correctly identify most true positive samples without making too many false positives.

## RESULTS

779 brain injury subjects and healthy controls enrolled during the study period and 331 were determined to have at least one valid biomarker result within 32 hours of injury. Of these eligible subjects, 21 expired during their initial admission with a final diagnosis of brain death. One subject was excluded due to invalid results on all three of the biomarkers of interest, GFAP, UCH-L1, and S100b, and another who suffered a gunshot wound to the head was excluded on the basis of penetrating brain injury. The remaining 19 brain death subjects (13 male, 6 female), had a mean (SD) age of 58.6 (17.6) with a range of 24-87. For comparative purposes, 51 isolated traumatic brain injury subjects with at least one valid biomarker result within 32 hours of injury were identified and further categorized as CT-positive (CTp) and CT-negative (CT-n) cohorts using the Marshall Classification (Marshall et al., 1992). 21 CTp subjects (15 male, 6 female) had a mean (SD) age of 56.2 (16.8) with a range of 31-85 and 30 CTn subjects (21 male, 9 female) had a mean (SD) age of 50.4 (25.5) with a range of 4-95. 15 non-trauma subjects that survived their initial admission (8 male, 7 female) had a mean (SD) age of 53.8 (15.4) with a range of 18-86. Additionally, 41 healthy control subjects (CTL) (11 male, 30 female) had a mean (SD) age of 37.1 (14.0) with a range of 16-65. Characteristics of all cohorts are displayed in Table 1.

The highest GCS scores were observed in the CTn group with 93% having a high GCS (13-15, N=28), while 95% (N=18) of the BD subjects had low GCS scores (8 or less) at the time of arrival to the emergency department. 62% (N=13) of CTp subjects had a high GCS (13-15) while the non trauma group had only 40% (N=6) with a high GCS and the majority (53%, N=8) had a low GCS. Loss of consciousness duration showed a similar trend with 50% (N=15) of CTn subjects having no loss of consciousness, while only 33% (N=5) of NT subjects had no LOC. 29% (N=6) of CTp had a loss of consciousness duration of 0 - 30 minutes, while most (74%, N=14) BD patients experienced greater than 24 hours of loss of consciousness prior to brain death diagnosis. Unknown loss of consciousness duration (unable to be obtained) remained a significant portion of all groups with 48% (N=10) of CTp and 53% (N=8) of non trauma patients having unknown loss of consciousness duration. A head CT was obtained for all brain injury patients except for one patient, who was in the brain death cohort. Mean (range) Marshall score for groups were 3.1 (1 - 6) for brain death subjects, 1 (1-1) for CTn, 2.14 (2 - 4) for CTp, and 2.47 (1 - 6) for the non trauma group. For the brain death subjects, the most prevalent mechanism of injury was found down (FD, 53%, N=10), followed by cardio-pulmonary arrest (CA/RA, 37%, N=7) and high velocity trauma/presumed diffuse axonal injury (11%, N=2).

Measures of central tendency (mean, median), dispersion (standard deviation (SD), interquartile range [25-75%] (IQR)) for the time differences from reported time of injury to blood sample collection for each of the four subject groups were comparable. The mean (SD | median | IQR) time of injury to blood draw was 720.5 minutes (543.4 | 562 | 270-1,227) for brain death, 671.8 minutes (529.9 | 465 | 234-1,284) for CTp, 581.7 minutes (598.4 | 295 | 140.75-1,214.25) for CTn, and 723.6 minutes (624.76 | 430 | 200-1,348) for non trauma. Within the brain death cohort, the mean (SD | median | IQR) time of injury to blood sample collection for each mechanism of injury was 8631 minutes (547.6 | 821 | 312-1,373) for FD, 505.1 minutes (501.8 | 320 | 205-525) for CA/RA, and 911.2 minutes (507.1 | 1,053 | 930-1,172) for diffuse axonal injury.

The number of samples and mean serum biomarker concentration for each patient cohort are reported in table 3. There were less patients overall for S100B samples. Serum concentrations of GFAP and UCH-L1 were found to be significantly different for each cohort. Detailed information about the total number of UCH-L1, GFAP, and S100B samples with detailed statistics and pg/mL concentrations for each patient cohort can be found in supplemental materials and figure 2. GFAP and UCH-L1 log concentrations were combined and evaluated as predictors of brain death using machine learning with Support Vector Machine (SVM) as shown in Table 4. Log-transformed concentrations of GFAP and UCH-L1 for each sample within the four subject groups and controls were plotted and shown in Figure 2 with contour lines. The area under the curve (AUC) for the brain death cohort was 0.95 (average precision of 0.85) compared to all others (CTL, non trauma, CTn, CTp); 0.94 [average precision of 0.85]) compared to all brain injury groups (non trauma, CTn, CTp); and 0.91 [average precision of 0.88]) compared to non trauma and CTp (Table 4). When the brain death group is delineated by mechanism of injury, as plotted in Figure 2, combinations of GFAP and UCH-L1 can predict brain death via CA/RA compared to brain death by FD or diffuse axonal injury with an AUC of 0.96 (average precision of 0.96, Table 4). The inclusion of the automated CT analysis (Blast-CT) into the multimodal algorithm improved the ability to differentiate brain death due to cardiac arrest from found down and diffuse axonal injury etiology.

**Figure 1:**
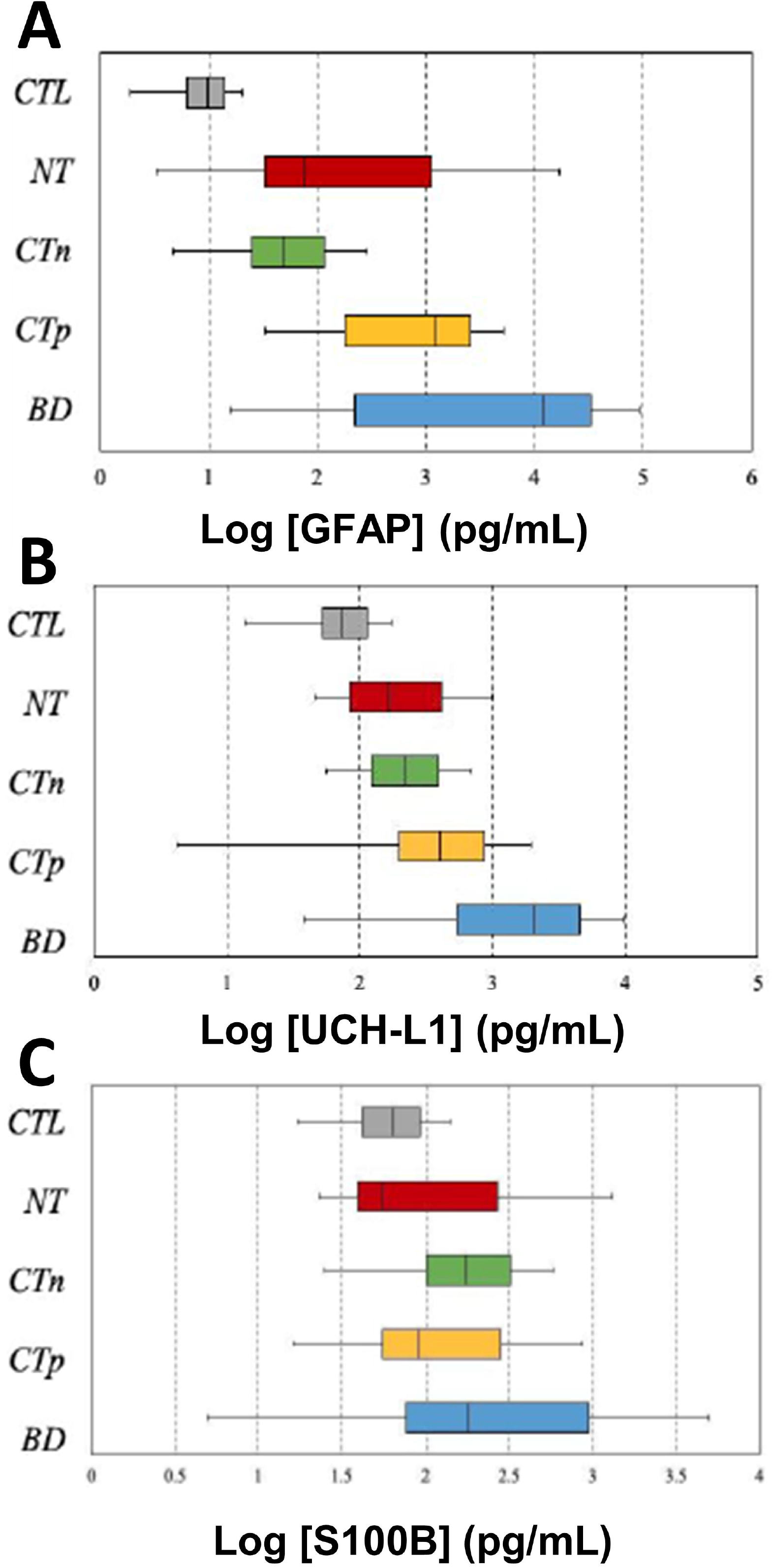
Boxplots of Log Concentrations of GFAP, UCH-L1 and S100B for Subject Groups and Controls. Concentrations of biomarkers are in picograms per milliliter (pg/mL). Values were log-transformed prior to plotting. Gray represents controls, red represents non-trauma, green represents CT-negative traumatic brain injury, yellow represents CT-positive traumatic brain injury, and blue represents brain death. [A] GFAP [B] UCH-L1 [C] S100B

**Figure 2:**
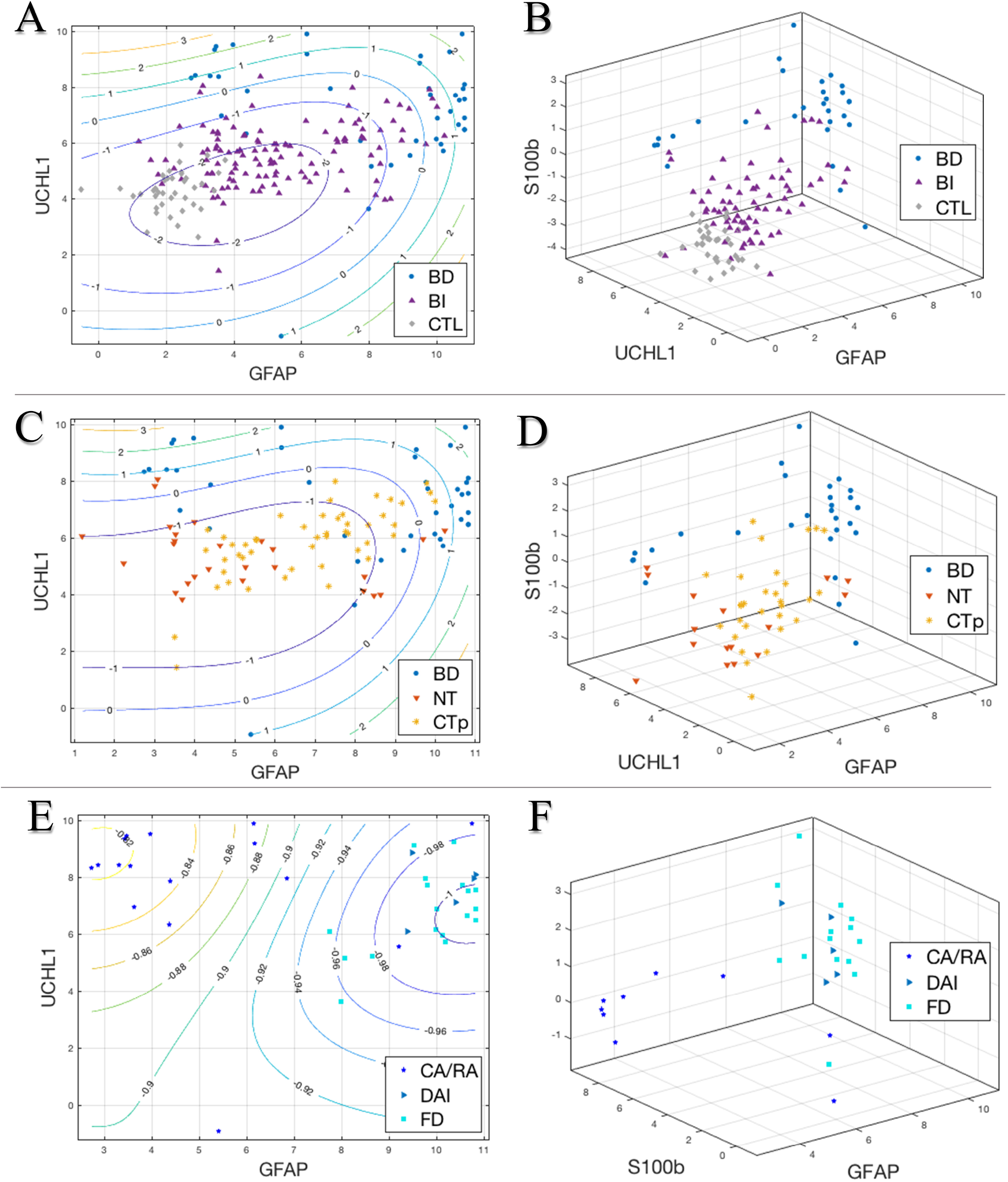
Contour and 3D scatterplots of Log Concentrations (pg/mL) of GFAP, UCH-L1, and S100b for comparison groups. [A] Contour scatterplot of log UCH-L1 and GFAP concentrations for brain death (BD) subjects, brain injury (BI) subjects which includes CT positive traumatic brain injury, CT negative traumatic brain injury, and non-trauma, and control (CTL) subjects. [B] 3D scatterplot of log S100b, UCH-L1, and GFAP concentrations for BD, BI, and CTL subjects. [C] Contour scatterplot of log UCH-L1 and GFAP concentrations for brain death (BD) subjects, non-trauma (NT) cohort from BI subjects which includes spontaneous intraparenchymal hemorrhages and hypoxic injury, and CT positive traumatic brain injury (CTp) subjects. [D] 3D scatterplot of log S100b, UCH-L1, and GFAP concentrations for BD, NT, and CTp subjects. [E] Contour scatterplot of log UCH-L1 and GFAP concentrations for brain death cohorts: cardiopulmonary arrest (CA/RA), diffuse axonal injury (DAI), and found down (FD) subjects. [F] 3D scatterplot of log S100b, UCH-L1, and GFAP concentrations for CA/RA, DAI, and FD subjects.

The ability of the combination of all 3 biomarkers to predict brain death was also assessed. Log-transformed concentrations of UCH-L1, GFAP, and S100B for each sample within the four subject groups and controls are shown in Figure 2. The AUC for UCH-L1, GFAP, and S100B combination in brain death was 0.98 (average precision of 0.92) compared to all others (CTL, non trauma, CTn, CTp); 0.96 (average precision of 0.91) compared to all brain injury groups (non trauma, CTn, CTp); and 0.93 (average precision of 0.91) when compared to non trauma and CTp (Table 5). The three biomarker combination was also able to distinguish CA/RA brain death from combined non trauma and diffuse axonal injury brain death with an AUC of 0.98 (average precision of 0.97, Table 4, Figure 2). Validation of the machine learning algorithm was assessed using two different measures of area under the curve and average precision which as shown in Table 4 and 5. The inclusion of the automated CT analysis (Blast-CT) into the multimodal algorithm with all three serum biomarker did not improve the ability to differentiate brain death due to cardiac arrest from found down and diffuse axonal injury etiology.

**Table 5.** SVM prediction accuracy from three serum biomarkers (GFAP, UCH-L1, and S100B) and inclusion of multimodal prediction with Blast-CT output.

| Comparison Groups<br>n = 137 samples | GFAP, UCH-L1, S100b |  | GFAP, UCH-L1, S100b, Blast-CT |  |
| --- | --- | --- | --- | --- |
|  | AUC | AP | AUC | AP |
| **BD vs. CTL, NT, CTn, CTp** | 0.98 | 0.92 | 0.96 | 0.89 |
| **BD vs. NT, CTn, CTp** | 0.96 | 0.91 | 0.94 | 0.89 |
| **BD vs. NT, CTp** | 0.93 | 0.91 | 0.93 | 0.90 |
| **CA vs. FD, DAI** | 0.98 | 0.97 | 0.98 | 0.97 |

## DISCUSSION

Brain injury is unlike injury to other organ systems in the body, because there are a lack of objective measures to classify the nature and severity of the problem. Without such measures, it is difficult to test therapeutics and prognosticate accurately (Rodriguez-Rodriguez et al., 2016). The aim of this study was to determine whether or not brain death could be predicted by a machine learning analysis of objective measures including algorithmic CT scan analysis and serum levels of GFAP, UCH-L1 and S100B following traumatic and atraumatic head injury. Our current ability to prognosticate neurologic outcome following cardiac arrest is limited and, similarly, accurate prognostication following a traumatic brain injury is difficult (Rodriguez-Rodriguez et al., 2016). Serum biomarkers have been investigated as a means to diagnose, risk-stratify and predict unfavorable outcome in both traumatic and non-traumatic brain injury patients, but few studies have used clinically-confirmed brain death as the main outcome measure (Egea-Guerrero et al., 2013; Wang et al., 2018). In the present study, biomarker concentrations within the first 32 hours post injury were evaluated for their ability to distinguish patients who eventually suffered brain death from those who survived. Further stratification in future work may enable even more precise prediction of deficit versus permanent disability (Geocadin et al., 2019). Biomarkers may identify patients for whom continuation of intensive life-sustaining efforts may prove futile, or conversely, identify patients for whom early induction of intensive therapy may provide maximal utility and offer improved outcomes (Okonkwo et al., 2020).

The findings of this study indicate that serum concentrations of three markers and CT scan analyses are different in subjects who died of brain death as a result of traumatic or atraumatic brain injury when compared to those who survived a traumatic brain injury with positive head CT, traumatic brain injury with negative head CT, non-traumatic brain injury, and controls (Figure 1). These findings are consistent with numerous studies that have found a strong association between GFAP and UCH-L1 levels in the acute post-traumatic brain injury setting and unfavorable outcome including death as measured by the Glasgow Outcome Scale (GOS) or extended GOS (GOSE) at 6 months post-injury (Di Battista et al., 2015; Mondello et al., 2016), as well as survival vs non-survival (Brophy et al., 2011; Mondello et al., 2012). Moreover, GFAP elevations have previously been shown to predict neurologic outcome following cardiac arrest (Larsson et al., 2014; Helwig et al., 2017). This study is among the first to demonstrate the utility of GFAP and UCH-L1 in predicting clinical brain death as opposed to simply predicting mortality.

Combinations of CT analysis plus two biomarkers (GFAP and UCH-L1, Table 4) and three biomarkers (GFAP, UCH-L1 and S100B, Table 5) were highly predictive of brain death with AUCs greater than 0.90 when compared to all others (CTL, non trauma, CTn, CTp; AUC = 0.95 and 0.98, AP = 0.85 and 0.92, respectively), brain injury groups (non trauma, CTn, CTp; AUC = 0.94 and 0.96, AP = 0.85 and 0.91, respectively), and non trauma and CTp (AUC = 0.91 and 0.93, AP = 0.88 and 0.91, respectively) (Table 4 &5). These AUC values were supported by high average precision (AP) for all comparisons. The inclusion of S100B to the model increased the AUC slightly over the two biomarker combination, indicating that S100B adds predictive value, despite the lack of significant difference in mean concentration compared to individual brain injury groups. As expected, the AUC is highest when compared to all others, which includes both brain injury groups and healthy controls. The AUC remains high even when comparing brain death only to non trauma and CTp, which is the most clinically relevant distinction.

Among our 19 brain death subjects, two suffered a high velocity traumatic brain injury consistent with diffuse axonal injury, seven suffered CA/RA and the remaining 10 were found down with unclear or mixed etiology brain injuries. Interestingly, our models with two and three biomarker combinations can distinguish between patients who suffered brain death via cardiac arrest (CA) as compared to a combined cohort of brain death from unknown/found down injury and diffuse axonal injury with an AUC of 0.96 and 0.98, respectively (Figure 2, Table 4). Of note, this AUC is higher than when comparing brain death as a whole to all others (CTL, non trauma, CTn, CTp; 3 biomarker AUC = 0.95) when only GFAP and UCH-L1 are used to predict brain death. Inclusion of the automated CT analysis improved the etiology classification of brain death subjects from AUC of 0.96 to 0.99. Blast-CT aided in classification and validation of the serum biomarker algorithm. It did not improve the prediction accuracy when all three serum biomarkers were included. Expanding our analysis with larger and more diverse datasets may help differentiate how the inclusion of lesion volume and type could be beneficial in a multimodal biomarker model.

As observed in Figure 2B, the profile of three serum markers clearly distinguish the brain death cohort from all other subjects. Figure 2D demonstrates that the CT positive cohort has a larger range of concentrations but is distinguishable between brain death and non trauma cohorts. Further, Figure 2F depicts a clear separation of brain death patients who suffered cardiac or respiratory arrest as compared to those found down with unknown injury and diffuse axonal injury. Our model represents a novel method to differentiate presumed diffuse axonal injury and FD patients from those who suffered cardiac or respiratory arrest (CA/RA), which is of particular importance in situations when the underlying neural insult is unknown or ambiguous. We hypothesize that the observed differences are a result of the dichotomous pathophysiology between traumatic and anoxic brain injuries. By comparing mechanism of injury within the brain death group, we avoid some of the variability in severity that was observed within the brain injury (CTp, CTn, and non trauma) cohorts. There are several limitations to this study, and the first of these is that the dataset consists of a relatively small number of patients. While the number appeared to be sufficient to develop the machine learning algorithm (table 4 and 5), it was insufficient to enable analysis on a validation set of data. We propose future work in which the model is tested on larger data sets such as that from the transforming research and clinical knowledge in traumatic brain injury initiative (TRACK-TBI) (Yue et al., 2013) or (CENTRE TBI) (Maas et al., 2015) datasets.

An additional caveat is that while objective measures have the potential to reduce biases in care, they need to be demonstrated to not perpetuate or exacerbate those biases. Automated analysis of CT scans is likely to be objective regardless of patient’s race, sex, gender, culture, education level or financial status, but whether or not a CT scan is obtained in the first place is almost certainly a function of these and other biases. Serum markers do show differences between races, with S100B having the most marked differences. Additional study will be needed to validate the use of serum markers for prognostication and classification of brain injury in diverse populations. And again, the decision to obtain serum markers in the first place remains a function of many factors potentially subject to bias.

We conclude that algorithmic CT analysis and serum levels of GFAP, UCH-L1 and S100B drawn within 32 hours post traumatic or non-traumatic brain injury can accurately identify patients who are at significant risk of developing brain death, highlighting the potential utility of our model to inform a challenging prognosis. Furthermore, these levels can also categorize brain death subjects by mechanism of injury into either cardiac/respiratory arrest or diffuse axonal injury. Classifying the nature of injury may be a helpful first step toward development of therapeutics to treat the consequences of injury.

## Data Availability

Data will be made available upon reasonable request.

## ETHICS STATEMENT

This study involving human participants were received and approved by the Human Subjects Research Committee and Institutional Review Board. The participants provided their informed written consent to participate in the clinical trial.

## DATA AVAILABILITY

Data will be made available upon reasonable request.

## AUTHOR CONTRIBUTIONS

US provided all oversight for the study. DR, RK, and US designed the study. ZL, BS, DR, BS, and TS were involved in data collection, data analysis, and manuscript writing. RK was involved in investigation and supervision of data analysis. All authors contributed to and approved the final manuscript.

## ACKNOWLEDGMENTS

We would like to thank the patients and controls who participated in the study, as well as the Minnesota Office of Higher Education and Abbott Labs for providing funding and resources for these studies.

## CONTRIBUTION TO THE FIELD STATEMENT

Assessment of the severity of brain injury is currently performed clinically and based on history and physical examination, along with radiologist interpretation of a CT scan. Such assessments are confounded by intoxicants, comorbidities and variability in clinical examination skills, along with examiner bias. Serum markers have recently been shown to predict a positive head CT scan and correlate with severity of brain injury. Algorithmic CT scan analysis demonstrates correlation with radiologist interpretation of scans. To date, no one has investigated whether these objective measures can predict brain death or determine its etiology. S100B, UCH-L1 and GFAP serum concentrations were collected within 32 hours following injury, in combination with algorithmic CT scan analysis, can be used to predict brain death in a group of trauma patients and classify its etiology in a binary fashion. This study represents an important addition to the literature because objective measures, if they can be demonstrated to be free of bias, have the potential to increase efficiency, reduce cost and reduce subjective influences in care of brain injured patients. Advances in the rapid classification and stratification of brain injury will ultimately enable testing and development of point of care therapeutics and improve patient outcomes

## REFERENCES

Brophy, G.M., Mondello, S., Papa, L., Robicsek, S.A., Gabrielli, A., Tepas, J., 3rd, et al. (2011). Biokinetic analysis of ubiquitin C-terminal hydrolase-L1 (UCH-L1) in severe traumatic brain injury patient biofluids. J Neurotrauma 28(6), 861–870. doi: 10.1089/neu.2010.1564.

Corinna Cortes, V.V. (1995). Support-vector networks. Machine Learning (20), 273–297.

Di Battista, A.P., Buonora, J.E., Rhind, S.G., Hutchison, M.G., Baker, A.J., Rizoli, S.B., et al. (2015). Blood Biomarkers in Moderate-To-Severe Traumatic Brain Injury: Potential Utility of a Multi-Marker Approach in Characterizing Outcome. Front Neurol 6, 110. doi: 10.3389/fneur.2015.00110.

Diaz-Arrastia, R., Wang, K.K., Papa, L., Sorani, M.D., Yue, J.K., Puccio, A.M., et al. (2014). Acute biomarkers of traumatic brain injury: relationship between plasma levels of ubiquitin C-terminal hydrolase-L1 and glial fibrillary acidic protein. J Neurotrauma 31(1), 19–25. doi: 10.1089/neu.2013.3040.

Egea-Guerrero, J.J., Murillo-Cabezas, F., Gordillo-Escobar, E., Rodriguez-Rodriguez, A., Enamorado-Enamorado, J., Revuelto-Rey, J., et al. (2013). S100B protein may detect brain death development after severe traumatic brain injury. J Neurotrauma 30(20), 1762–1769. doi: 10.1089/neu.2012.2606.

Faul, M., and Coronado, V. (2015). Epidemiology of traumatic brain injury. Handb Clin Neurol 127, 3–13. doi: 10.1016/B978-0-444-52892-6.00001-5.

Fawcett, T. (2006). An introduction to ROC analysis. Pattern recognition letters. 27(8), 861–874.

Geocadin, R.G., Callaway, C.W., Fink, E.L., Golan, E., Greer, D.M., Ko, N.U., et al. (2019). Standards for Studies of Neurological Prognostication in Comatose Survivors of Cardiac Arrest: A Scientific Statement From the American Heart Association. Circulation 140(9), e517–e542. doi: 10.1161/CIR.0000000000000702.

Greer, D.M., Shemie, S.D., Lewis, A., Torrance, S., Varelas, P., Goldenberg, F.D., et al. (2020). Determination of Brain Death/Death by Neurologic Criteria: The World Brain Death Project. JAMA 324(11), 1078–1097. doi: 10.1001/jama.2020.11586.

Helwig, K., Seeger, F., Holschermann, H., Lischke, V., Gerriets, T., Niessner, M., et al. (2017). Elevated Serum Glial Fibrillary Acidic Protein (GFAP) is Associated with Poor Functional Outcome After Cardiopulmonary Resuscitation. Neurocrit Care 27(1), 68–74. doi: 10.1007/s12028-016-0371-6.

Larsson, I.M., Wallin, E., Kristofferzon, M.L., Niessner, M., Zetterberg, H., and Rubertsson, S. (2014). Post-cardiac arrest serum levels of glial fibrillary acidic protein for predicting neurological outcome. Resuscitation 85(12), 1654–1661. doi: 10.1016/j.resuscitation.2014.09.007.

Lingsma, H.F., Roozenbeek, B., Steyerberg, E.W., Murray, G.D., and Maas, A.I. (2010). Early prognosis in traumatic brain injury: from prophecies to predictions. Lancet Neurol 9(5), 543–554. doi: 10.1016/S1474-4422(10)70065-X.

Maas, A.I., Menon, D.K., Steyerberg, E.W., Citerio, G., Lecky, F., Manley, G.T., et al. (2015). Collaborative European NeuroTrauma Effectiveness Research in Traumatic Brain Injury (CENTER-TBI): a prospective longitudinal observational study. Neurosurgery 76(1), 67–80. doi: 10.1227/NEU.0000000000000575.

Mahan, M.Y., Thorpe, M., Ahmadi, A., Abdallah, T., Casey, H., Sturtevant, D., et al. (2019). Glial Fibrillary Acidic Protein (GFAP) Outperforms S100 Calcium-Binding Protein B (S100B) and Ubiquitin C-Terminal Hydrolase L1 (UCH-L1) as Predictor for Positive Computed Tomography of the Head in Trauma Subjects. World neurosurgery 128, e434–e444. doi: 10.1016/j.wneu.2019.04.170.

Marshall, L.F., Marshall, S.B., Klauber, M.R., Van Berkum Clark, M., Eisenberg, H., Jane, J.A., et al. (1992). The diagnosis of head injury requires a classification based on computed axial tomography. J Neurotrauma 9 Suppl 1, S287–292.

Mesfin, F.B., Gupta, N., Hays Shapshak, A., and Taylor, R.S. (2020). “Diffuse Axonal Injury,” in StatPearls. (Treasure Island (FL)).

Mondello, S., Kobeissy, F., Vestri, A., Hayes, R.L., Kochanek, P.M., and Berger, R.P. (2016). Serum Concentrations of Ubiquitin C-Terminal Hydrolase-L1 and Glial Fibrillary Acidic Protein after Pediatric Traumatic Brain Injury. Scientific reports 6, 28203. doi: 10.1038/srep28203.

Mondello, S., Linnet, A., Buki, A., Robicsek, S., Gabrielli, A., Tepas, J., et al. (2012). Clinical utility of serum levels of ubiquitin C-terminal hydrolase as a biomarker for severe traumatic brain injury. Neurosurgery 70(3), 666–675. doi: 10.1227/NEU.0b013e318236a809.

Mussack, T., Biberthaler, P., Kanz, K.G., Wiedemann, E., Gippner-Steppert, C., Mutschler, W., et al. (2002). Serum S-100B and interleukin-8 as predictive markers for comparative neurologic outcome analysis of patients after cardiac arrest and severe traumatic brain injury. Crit Care Med 30(12), 2669–2674. doi: 10.1097/00003246-200212000-00010.

Nguyen, T.V., Hayes, M., Zbesko, J.C., Frye, J.B., Congrove, N.R., Belichenko, N.P., et al. (2018). Alzheimer’s associated amyloid and tau deposition co-localizes with a homeostatic myelin repair pathway in two mouse models of post-stroke mixed dementia. Acta Neuropathol Commun 6(1), 100. doi: 10.1186/s40478-018-0603-4.

Okonkwo, D.O., Puffer, R.C., Puccio, A.M., Yuh, E.L., Yue, J.K., Diaz-Arrastia, R., et al. (2020). Point-of-Care Platform Blood Biomarker Testing of Glial Fibrillary Acidic Protein versus S100 Calcium-Binding Protein B for Prediction of Traumatic Brain Injuries: A Transforming Research and Clinical Knowledge in Traumatic Brain Injury Study. J Neurotrauma 37(23), 2460–2467. doi: 10.1089/neu.2020.7140.

Papa, L., Brophy, G.M., Welch, R.D., Lewis, L.M., Braga, C.F., Tan, C.N., et al. (2016). Time Course and Diagnostic Accuracy of Glial and Neuronal Blood Biomarkers GFAP and UCH-L1 in a Large Cohort of Trauma Patients With and Without Mild Traumatic Brain Injury. JAMA neurology 73(5), 551–560. doi: 10.1001/jamaneurol.2016.0039.

Rodriguez-Rodriguez, A., Egea-Guerrero, J.J., Gordillo-Escobar, E., Enamorado-Enamorado, J., Hernandez-Garcia, C., Ruiz de Azua-Lopez, Z., et al. (2016). S100B and Neuron-Specific Enolase as mortality predictors in patients with severe traumatic brain injury. Neurol Res 38(2), 130–137. doi: 10.1080/01616412.2016.1144410.

Rosenfeld, J.V., Maas, A.I., Bragge, P., Morganti-Kossmann, M.C., Manley, G.T., and Gruen, R.L. (2012). Early management of severe traumatic brain injury. Lancet 380(9847), 1088–1098. doi: 10.1016/S0140-6736(12)60864-2.

Wang, K.K., Yang, Z., Zhu, T., Shi, Y., Rubenstein, R., Tyndall, J.A., et al. (2018). An update on diagnostic and prognostic biomarkers for traumatic brain injury. Expert Rev Mol Diagn 18(2), 165–180. doi: 10.1080/14737159.2018.1428089.

Yue, J.K., Vassar, M.J., Lingsma, H.F., Cooper, S.R., Okonkwo, D.O., Valadka, A.B., et al. (2013). Transforming research and clinical knowledge in traumatic brain injury pilot: multicenter implementation of the common data elements for traumatic brain injury. J Neurotrauma 30(22), 1831–1844. doi: 10.1089/neu.2013.2970.

Zhang E. Z.Y. (2009). Average Precision. Encyclopedia of Database Systems. Springer, Boston, MA.

