## Supplemental Materials for "Machine Learning with Objective Serum Markers and Algorithmic Deep Learning CT Scan Analysis Predicts Brain Death Due to Trauma and Hypoxia"

*Study Setting and Population*

Subjects with a history of head trauma in the last six months, penetrating trauma, those actively participating in another clinical study, or with active psychiatric, neurological, and developmental disorders were excluded. Additionally, protected populations such as prisoners, homeless, and pregnant patients were not included in this study. All eligible subjects were between the ages of 4-100 years old with a suspected traumatic or non-traumatic brain injury, and at least one blood specimen collected within 32 hours of injury. Only blood specimens with valid GFAP, UCH-L1, and S100b biomarker concentrations were included. For comparison, healthy controls (CTL) were also identified and recruited by screening family members and visitors of subjects. Control subjects were excluded if they had suffered a head injury within 1 year of enrollment or were participating in an experimental drug or device trial.

*Key Outcome Measures*

Subjects that expired during their initial hospitalization as a result of either traumatic or non-traumatic brain injuries formed the brain death cohort. The diagnosis of brain death was made clinically by a care team independent of and blinded to the observational study in accordance with hospital guidelines. The brain death exam consisted of the absence of motor responses, loss of brainstem reflexes and apnea, in the absence of potentially confounding factors including certain physiological conditions and drug or chemical interactions (Wijdicks, 2001). At the discretion of the care team, additional confirmatory neuroimaging studies were performed including CT, MRI and nuclear scintigraphy cerebral blood flow. Confirmatory findings included absent cerebral blood flow on nuclear scintigraphy (Rizvi et al., 2018). Each BD subject was categorized based on mechanism of injury into either diffuse axonal injury, cardiac/respiratory arrest (CA/RA) or found down/unknown etiology (FD) cohorts.

The remaining subjects who survived to hospital discharge formed the brain injury group, which was subdivided into three groups based on mechanism of injury and clinical CT findings and Marshall Classification (Marshall et al., 1992) by clinical radiologists. Subjects with suspected head trauma were divided into CT-positive (CTp, Marshall score 2 - 6) or CT-negative (CTn, Marshall score 1). The non-trauma (NT) group was determined by the clinical care team to have a brain injury other than a TBI which included anoxic and hypoxic injuries such as cardiac or respiratory arrest (CA/RA), opioid overdose, spontaneous intraparenchymal hemorrhage, or who were otherwise found down (FD) with unclear or mixed etiology brain injuries.

Classification based on the integrated features of serum biomarkers and CT scan images was performed using a pre-trained deep learning model (blast-ct) based on a convolutional neural network (Monteiro et al., 2020). This algorithm automatically generates the voxel-wise segmentation of 4 types of lesions. Blast-CT was utilized by providing a new dataset of CT scans from serum biomarker matched patients. Blast-CT identified the regions of the four lesion-types and quantified their respective volumes by counting the total segmented voxels. The segmentation volumes were normalized by the total head volume in each scan.

*Blood Collection and Processing*

Serum (18 mL) and plasma (12 mL) samples were collected at the time of enrollment in serum-separating tubes (SST) and K2EDTA vacutainers, respectively. The first blood sample was collected at admission (0 hr), second drawn within 8 hours of the reported time of injury, and a third was drawn 12 - 32 hours post-injury. For the development of the machine learning algorithm capable of stratification by injury etiology, all available serum biomarker concentrations taken from the same blood draw the were utilized. Due to the nature of traumatic injuries many patients only had one or two blood draws taken. There was no correlation between injury etiology and number of blood draws taken. At each collection point, approximately 40 mL of blood was drawn and distributed evenly into SST and EDTA vacutainers. Both sample types were held at room temperature for between 30 minutes at 2 hours prior to centrifugation for 10 minutes at 2380 rpm. Processed samples were aliquoted and stored at -80°C prior to shipment on dry ice to Abbott Laboratories (Abbott Park, IL) for assay analysis. Serum samples were analyzed for UCH-L1, GFAP, and S100B. Results were reported in picograms/milliliter (pg/mL) for all biomarkers.

*Data Analysis*

Analysis and plotting was performed using Microsoft Excel 2016 (v16.0) and MATLAB (R2018b). Subject characteristics including demographic information, loss of consciousness duration, Glasgow Coma Score (GCS) score, percentage with clinical CT, Marshall score, length of hospital stay, and discharge location were described using descriptive statistics. Only samples with at least one valid UCH-L1, GFAP, or S100B biomarker result were included in the analysis. Cohort biomarker levels were compared to blood samples from healthy controls from a single blood draw. Biomarker concentrations were log-transformed and values below the detectable limit prior to transformation were assigned a value of zero, but included in the analysis. Biomarker levels found to be above the detectable limit (50,000 pg/ml for GFAP and 20,000 pg/ml for UCH-L1) were included in analysis as equal to the upper limit.

Descriptive statistics including mean, median, standard deviation, and interquartile range (IQR) were calculated where appropriate. Differences in mean raw concentrations between CTL, CTp, CTn, NT and BD groups at all timepoints measured within 32 hours were assessed using the t-test. Concentrations were log-transformed to achieve normality prior to plotting and ROC analysis.

*Machine Learning*

Support Vector Machine (SVM) (Corinna Cortes, 1995) was utilized for the classification of the patient samples in our prediction tasks. Among the supervised learning methods, SVM generalizes well on new test data, and the decision function of SVM is simply defined by a subset of training samples without requiring training with big data.

Support vector machine and kernels:

Given a set of training data
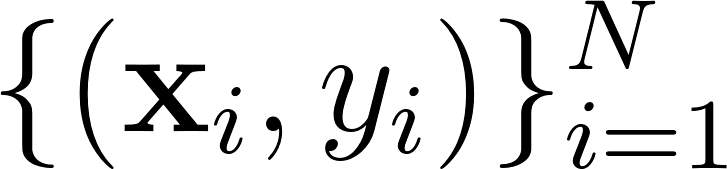
<https://www.codecogs.com/eqnedit.php?latex=%5C%7B%20(%5Cmathbf%7Bx%7D_i%2C%20y_i%20)%5C%7D%5EN_%7Bi%3D1%7D%250> with
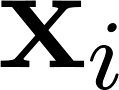
<https://www.codecogs.com/eqnedit.php?latex=%5Cmathbf%7Bx%7D_i%250> be the
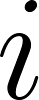
<https://www.codecogs.com/eqnedit.php?latex=i%250>-th sample vector and
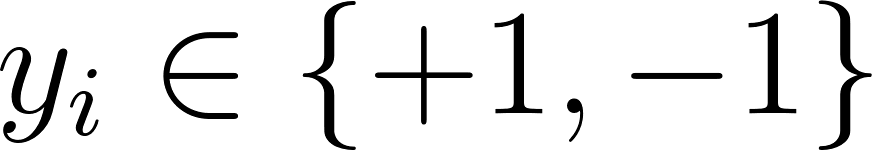
<https://www.codecogs.com/eqnedit.php?latex=y_i%20%5Cin%20%5C%7B%2B1%2C-1%5C%7D%250> as its label, the SVM outputs the score
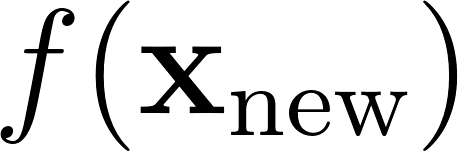
<https://www.codecogs.com/eqnedit.php?latex=f(%5Cmathbf%7Bx%7D_%5Ctext%7Bnew%7D)%250> of a new sample
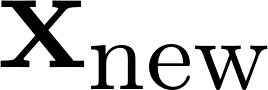
<https://www.codecogs.com/eqnedit.php?latex=%5Cmathbf%7Bx%7D_%7B%5Ctext%7Bnew%7D%7D%250> as

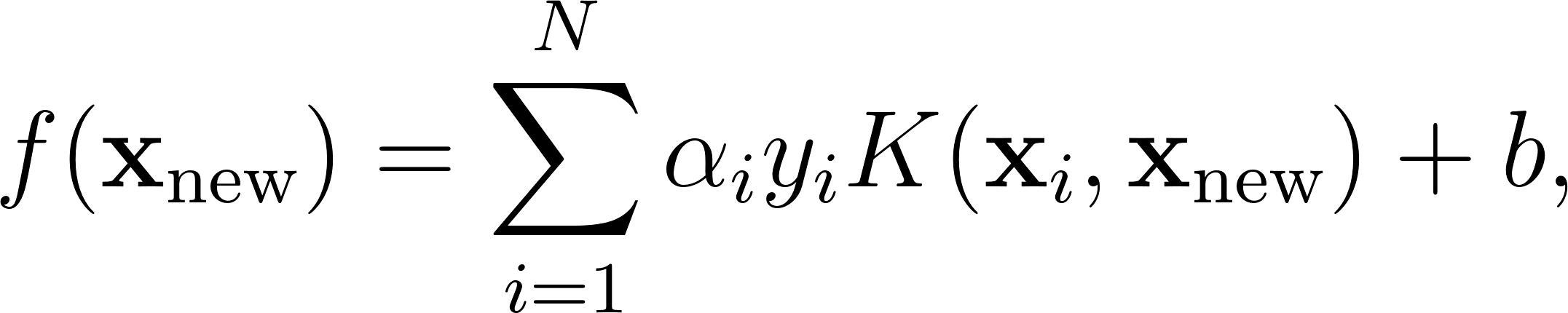
<https://www.codecogs.com/eqnedit.php?latex=f(%5Cmathbf%7Bx%7D_%7B%5Ctext%7Bnew%7D%7D)%20%3D%20%5Csum%5Climits_%7Bi%3D1%7D%5E%7BN%7D%20%5Calpha_i%20y_i%20K(%5Cmathbf%7Bx%7D_i%2C%20%5Cmathbf%7Bx%7D_%7B%5Ctext%7Bnew%7D%7D)%20%2B%20b%2C%250> (1)

where
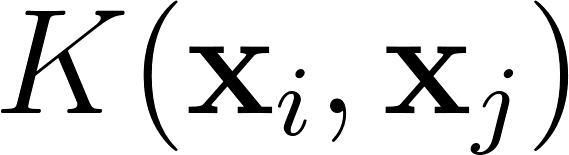
<https://www.codecogs.com/eqnedit.php?latex=K(%5Cmathbf%7Bx%7D_i%2C%5Cmathbf%7Bx%7D_j)%250> is the kernel function defined with a mapping of the original feature space into a transformed space for non-linear classification. We use the Gaussian kernel defined below as the non-linear kernel function:

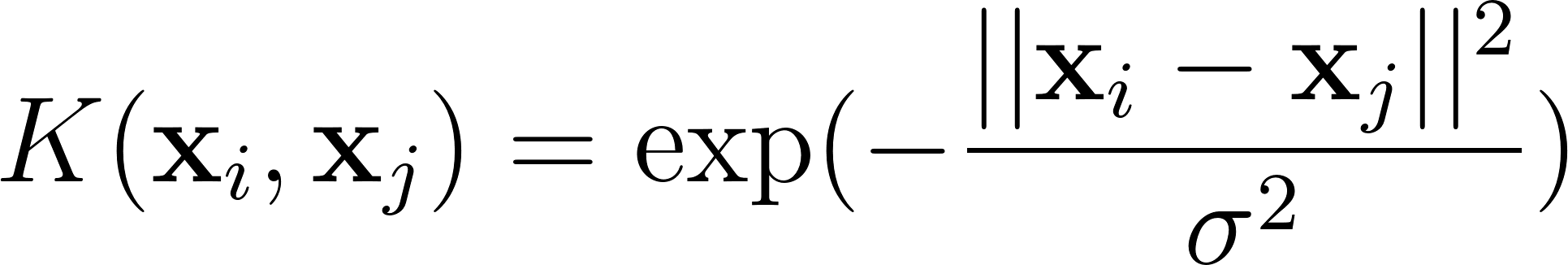
<https://www.codecogs.com/eqnedit.php?latex=K(%5Cmathbf%7Bx%7D_i%2C%20%5Cmathbf%7Bx%7D_j)%20%3D%20%5Ctext%7Bexp%7D%20(-%5Cfrac%7B%7C%7C%5Cmathbf%7Bx%7D_i%20-%20%5Cmathbf%7Bx%7D_j%7C%7C%5E2%7D%7B%5Csigma%5E2%7D)%250> (2)

The
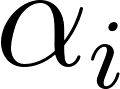
<https://www.codecogs.com/eqnedit.php?latex=%5Calpha_i%250> for
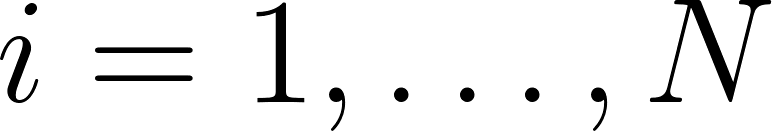
<https://www.codecogs.com/eqnedit.php?latex=i%3D1%2C%5Cdots%2C%20N%250> are obtained by solving the optimization problem

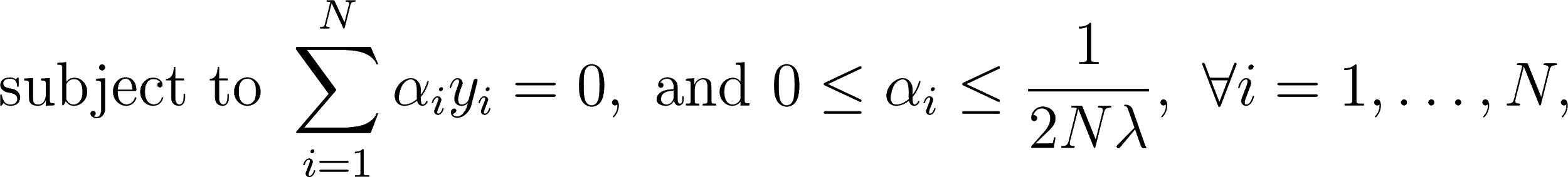

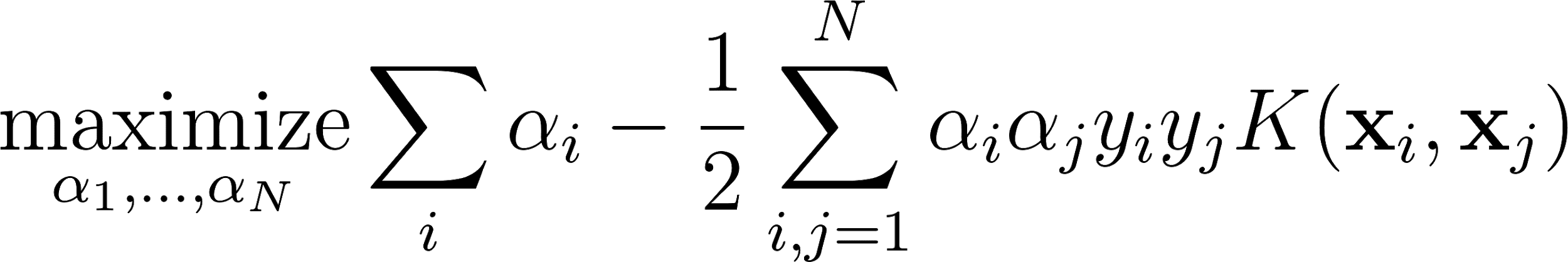
<https://www.codecogs.com/eqnedit.php?latex=%5Cmathop%7B%5Ctext%7Bmaximize%7D%7D_%7B%5Calpha_1%2C%20%5Cdots%2C%20%5Calpha_N%7D%20%5Csum_i%20%5Calpha_i%20-%20%5Cfrac%7B1%7D%7B2%7D%20%5Csum_%7Bi%2Cj%3D1%7D%5EN%20%20%5Calpha_i%20%5Calpha_j%20y_i%20y_j%20K(%5Cmathbf%7Bx%7D_i%2C%5Cmathbf%7Bx%7D_j)%250> (3)

(4)

where
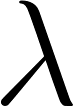
<https://www.codecogs.com/eqnedit.php?latex=%5Clambda%250> is a hyper-parameter.

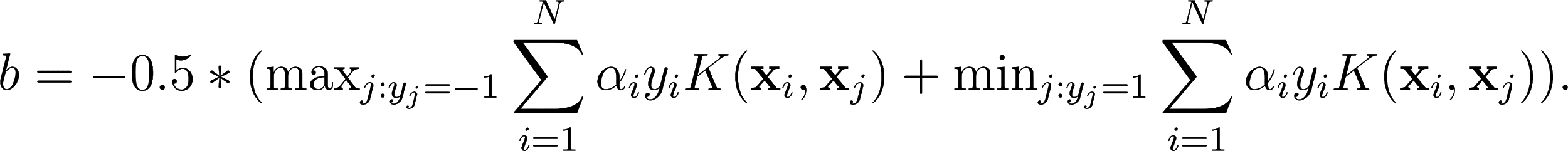
After obtaining all the
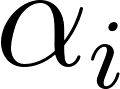
<https://www.codecogs.com/eqnedit.php?latex=%5Calpha_i%250>s, the scalar
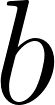
<https://www.codecogs.com/eqnedit.php?latex=b%250> is obtained as:

*(5)*

*Contour plot*

To generate the contour plots in Figure 2 A, C and E, we first generated a set of 2-D grid coordinates in the plane. Then, we predicted the scores of these 2-D grid coordinates using the SVM model trained using all the samples with the optimal parameters obtained in the leave-one-out cross-validation. Finally, we visualized the contours of the scores of the 2-D grid coordinates.

*GFAP, UCH-L1, and S100B biomarker supplemental information*

A total of 203 valid UCH-L1 samples were analyzed, 40 were in the brain death group, 46 in the CTp group, 49 in the CTn group, 27 in the non trauma group, and 41 in the control group. Mean (SD) time from injury to blood collection was similar for all groups with 731·6 minutes (545·6) for brain death, 688·0 minutes (534·7) for CTp, 581·7 minutes (598.4) for CTn, and 723·6 minutes (624·8) for non trauma cohorts. Brain death subjects had a median UCH-L1 concentration of 2,082.5 pg/mL (IQR: 537·5-4,506·3 pg/mL) compared to 395·9 pg/mL (IQR: 196·4-872·9 pg/mL) for CTp, 218·3 pg/mL (IQR: 122-388·2 pg/mL) for CTn, 164·5 pg/mL (IQR: 84·8-408 pg/mL) for non trauma cohort, and 73·7 pg/mL (IQR: 44·2-113·1 pg/mL) for CTL subjects (Figure 1). Mean concentrations of UCH-L1 in the brain death group were significantly higher than all other comparison groups and controls (p < 0·001; t-test). Of the brain death subjects, median (IQR) UCH-L1 concentrations were 775·6 pg/mL (411·95-2,082·5 pg/mL) for FD, 4,421·9 pg/mL (2,228·3-11,774·1 pg/mL) for CA/RA, and 2,830·4 pg/mL (1,245·8-3,304·1 pg/mL) for diffuse axonal injury. UCH-L1 concentrations are reported for each patient and each blood draw time point in Table 2.

203 valid GFAP samples were analyzed, 40 within the brain death group, 46 within the CTp group, 49 within the CTn group, 27 within the non trauma group, and 41 within the CTL group. Mean (SD) time from injury to blood collection was similar for all groups with 731·6 minutes (545·6) for brain death, 688·0 minutes (534·7) for CTp, 581·7 minutes (598.4) for CTn, and 723·6 minutes (624·8) for non trauma cohorts. Brain death subjects had a median GFAP concentration of 12,622.7 pg/mL (IQR: 185.7-34,339.1 pg/mL) compared to 1,230.6 pg/mL (IQR: 181·9-2,527·1 pg/mL) for CTp, 48.7 pg/mL (IQR: 25·1-118·2 pg/mL) for CTn, 77·7 pg/mL (IQR: 33·4-1,833·0 pg/mL) for non trauma, and 9·4 pg/mL (IQR: 5·8-13·3 pg/mL) for CTL subjects (Figure 1). Mean concentrations of GFAP in the brain death group were significantly higher than all other comparison groups and controls (p < 0·001; t-test). In the brain daeth cohort, median (IQR) concentrations of GFAP were 24,959·7 pg/mL (15,565·2-42,394·5 pg/mL) for FD, 65·75 pg/mL (31·9-472 pg/mL) for CA/RA, and 33,018·7 pg/mL (13,284·3-48,064·2 pg/mL) for diffuse axonal injury. GFAP concentrations are reported for each patient and each blood draw time point in Table 2

137 valid S100B samples were analyzed, 28 from the brain death group, 30 from the CTp group, 24 from the CTn group, 17 from the non trauma group, and 38 from the CTL group. Mean (SD) time from injury to blood collection was similar for all groups with 749·3 minutes (569·5) for brain death, 463·6 minutes (569·9) for CTn, 628·6 minutes (523·3) for CTp, and 733·0 minutes (635·4) for non trauma cohorts. Brain death subjects had a median S100B concentration of 180 pg/mL (IQR: 77·5-945 pg/mL) compared to 91 pg/mL (IQR: 57·8-281 pg/mL) for CTp, 177·5 pg/mL (IQR: 100·3-321·5 pg/mL) for CTn, 55 pg/mL (IQR: 39-269 pg/mL) for non trauma, and 63 pg/mL (IQR: 41·8-94 pg/mL) for CTLs (Figure 1). Mean concentrations of S100B were significantly higher in brain death subjects than controls (p < 0·001; t-test), but no significant differences were present between the brain death subjects and any of the comparison groups. Of the brain death subjects, median (IQR) concentration of S100B was 210 pg/mL (102·3-612·25 pg/mL) for FD, 55 pg/mL (52-563 pg/mL) for CA/RA, and 1,030 pg/mL (134-1,060 pg/mL) for diffuse axonal injury. S100B concentrations are reported for each patient and each blood draw time point in Table 2. GFAP and UCH-L1 serum concentrations were found to be significantly difference for brain death individually compared to non trauma, CTp, CTn. or uninjured controls with p-values well under 0.05 as shown in Table 3. There were less samples available with valid S100B concentrations. S100B only had statistically significant results for differentiating brain death from uninjured controls.

**Supplemental Materials References**

Corinna Cortes, V.V. (1995). Support-vector networks. *Machine Learning* (20)**,** 273-297.

Marshall, L.F., Marshall, S.B., Klauber, M.R., Van Berkum Clark, M., Eisenberg, H., Jane, J.A., et al. (1992). The diagnosis of head injury requires a classification based on computed axial tomography. *J Neurotrauma* 9 Suppl 1**,** S287-292.

Monteiro, M., Newcombe, V.F.J., Mathieu, F., Adatia, K., Kamnitsas, K., Ferrante, E., et al. (2020). Multiclass semantic segmentation and quantification of traumatic brain injury lesions on head CT using deep learning: an algorithm development and multicentre validation study. *Lancet Digit Health* 2(6)**,** e314-e322. doi: 10.1016/S2589-7500(20)30085-6.

Rizvi, T., Batchala, P., and Mukherjee, S. (2018). Brain Death: Diagnosis and Imaging Techniques. *Semin Ultrasound CT MR* 39(5)**,** 515-529. doi: 10.1053/j.sult.2018.01.006.

Wijdicks, E.F. (2001). The diagnosis of brain death. *N Engl J Med* 344(16)**,** 1215-1221. doi: 10.1056/NEJM200104193441606.
